# Family physicians supporting patients with palliative care needs within the Patient Medical Home in the community: An Appreciative Inquiry qualitative study

**DOI:** 10.1101/2021.01.04.21249226

**Authors:** Amy Tan, Ronald Spice, Aynharan Sinnarajah

## Abstract

**Objectives:** Canadians want to live and die in their home communities. Unfortunately, Canada has the highest proportion of deaths in acute care facilities as compared to other developed nations. This study aims to identify the essential components required to best support patients and families with palliative care needs in their communities to inform system changes and empower family physicians (FPs) in providing community-based palliative care for patients.

**Design:** Appreciative Inquiry (AI) methodology with individual interviews. Interview transcripts were analyzed iteratively for emerging themes and used to develop “possibility statements” to frame discussion in subsequent focus groups. A conceptual framework emerged to describe the “destiny” state as per AI methods.

**Setting:** Family physicians, palliative home care providers, patients and bereaved caregivers were recruited in the urban and surrounding rural health authority zones of Calgary, Canada.

**Participants:** 9 females and 9 males FPs (range of practice years 2-42) in interviews; 8 bereaved caregivers, 1 patient, 26 Palliative Home Care team members in focus groups. Interviews and focus groups were recorded digitally and transcribed with consent.

**Results:** The identified themes that transcended all three groups created the foundation for the conceptual framework. Enhanced communication and fostering team relationships between all care providers with the focus on the patient and caregivers was the cornerstone concept. The family physician/patient relationship must be protected and encouraged by all care providers, while more system flexibility is needed to respond more effectively to patients. These concepts must exist in the context that patients and caregivers need more education regarding the benefits of palliative care, while increasing public discourse about mortality.

**Conclusions:** Key areas were identified for how the patient’s team can work together effectively to improve the patient and caregiver palliative care journey in the community with the cornerstone element of building on the trusting FP-patient longitudinal relationship.

**Strengths and limitations of this study:**

- This study uses Appreciative Inquiry qualitative methods to identify the essential components required to best support patients and families with palliative care needs in their communities.
- Multiple perspectives recruited including: community-based family physicians, palliative home care clinicians, patients, and bereaved caregivers.
- Analysis was focused on the family physicians’ interviews to derive “possibility statements” and used to frame the focus groups with the other groups of participants.
- Generalisability may be limited due to the lack of diversity in participants recruited for the patients and bereaved caregivers in terms of ethnicity, age, and gender.
- Patients and caregivers may have been reluctant to volunteer for this study as it involved discussing palliative care.

## Background

Canadians want to live and die in their communities [1] as they are growing older and living longer with chronic conditions and multimorbidity. Unfortunately, Canada has the highest proportion of deaths in acute care facilities (62%) and higher mean per capita hospital expenditures as compared to other developed nations [2, 3] There is a disconnect. Although community-based primary palliative care (CPPC) is a core competency for Canadian family physicians (FPs) [4], less FPs self-identify as providing CPPC to their patients. 3 out of 5 primary care physicians in Canada report that they are unprepared to meet the palliative care needs of their patients.[1] Palliative care within primary care is also weakening in other developed nations in the world, including the UK, the Netherlands and Australia [5, 6, 7].

Palliative care is defined by the WHO as an approach to care that improves the quality of life of people with serious or life-limiting diseases, provides support to their families, and focuses on the assessment and management of physical symptoms and psychosocial and spiritual concerns.[8] Primary palliative care is defined as palliative care provided by “front-line” clinicians, including primary care physicians and oncologists, that is integrated into the care of patients in a patient-centred approach [6, 9]. Given that family physicians in Canada practice holistic patient-centred care where their patients live, a purposeful integration of CPPC within family medicine in Canada is both appropriate and essential for high quality and efficient primary care.

It is not known why there is a discrepancy between the increasing need for Canadian FPs to support their patients with palliative care needs, and the decreasing availability of FPs to provide such care. Consequently, it is unclear as to how FPs can be better supported in their practices to provide for the care of palliative patients in their communities.

To understand the impact of low community-based FP involvement in palliative care, one must determine what is required to better prepare and empower FPs for the increasing palliative care needs of an aging population with multi-morbidity [6]. Improved FP involvement would also allow patients to benefit from an earlier palliative approach and more effective chronic disease management [6, 10].

This study aims to explore and describe the current landscape of FPs’ involvement in providing CPPC to patients within the Patient Medical Home (PMH) [11] in a metropolitan city and surrounding rural communities in Southern Alberta, Canada. We have included the perspectives of palliative home care providers who are key partners in the delivery of community-based palliative care. Further, we have explored the perspectives of the recipients of the care: patients with palliative care needs and their unpaid caregivers. These perspectives were explored to identify the essential components required to best support patients and families with palliative care needs in their communities throughout their illness trajectories to inform system changes and empower FPs in providing CPPC for patients.

## METHODS

A qualitative method with an Appreciative Inquiry (AI) approach was utilized for this study. The design, data collection and analysis align with the Standards for Reporting Qualitative Research (SRQR) [12] The AI approach encourages a strengths-based, forward-looking approach that focuses on positive changes and solutions to maintain and build on successes encountered by clinicians who are supporting patients and their families with community-based palliative care needs.[13-16]. Participants were asked to “discover”, “dream”, and “design”[13] their ideal environment to best support their patients with palliative care needs, and discuss their role and needs with this environment. Patients and caregivers were asked to determine what they would need to better support their loved ones if they were to advise others to achieve a better palliative care experience. Their responses informed interviews and focus groups toto identify and build on what works well within organizations, stimulates innovation and creates a future “destiny”, rather than examining only what is broken [13-16].

This study received ethical approval from the Conjoint Health Research Ethics Board (CHREB) at the University of Calgary, Canada.

## STUDY SETTING AND SAMPLE

Recruitment of the FPs were recruited from Calgary area Primary Care Networks (PCN) through advertising in PCN newsletters and used PCN email distribution lists to find interested family physicians. PCNs are geographically located networks of FPs and interdisciplinary primary care clinicians working in team-based environments to improve primary care access and care for patients.[17]). Contacting the different PCNs allowed for purposeful [18] sampling of family physicians from both urban and rural regions within the Calgary health zone. The study was advertised in the electronic newsletters of the Alberta Health Services’ Departments of Family Medicine and Rural Medicine and study information was faxed and emailed to every clinic. Snowball sampling was also used by providing each participant with contact information to pass along to colleagues who may be interested in participating in the study. Theoretical sampling[19] was used to guide data collection for further insights to support the team’s evolving understanding of emerging concepts. Informed consent was obtained from each participant at the beginning of the interview.

Theoretical sampling was also used in recruiting Palliative Home Care (PHC) clinicians to better understand the perspectives of palliative providers who work with FPs. Two PHC teams from different quadrants of the city, each with its own unique patient demographics, volunteered to participate. Patients and caregivers of patients with palliative care needs were recruited through the Alberta Health Services’ Patient and Family Advisory Group, a group of public volunteers with a variety of healthcare patient experiences [20], who provided input on the patient and caregiver experience with receiving palliative care.

## DATA COLLECTION

The FPs participated in individual semi-structured interviews with a research assistant conducted either in person, by telephone, or through Skype for Business (tm) depending on the participant’s preference. A semi-structured interview guide was developed using AI principles to assess and explore the current clinical landscape regarding family medicine and the provision of CPPC. Demographic information including gender, years of practice, additional palliative care training, and country of medical school was collected (Table 1).

**Table 1:** **Demographics Collected for Family Physicians, Patients and Caregivers, and Palliative Home Care Team Members**

| # Family Physicians Interviewed | Gender | Location of Medical School | Additional Formal Palliative Care Training | Duration of FM practice (years) | Interview Length Range (minutes) | Rural or Urban practice experience | Practice setting in addition to FM clinic |
| --- | --- | --- | --- | --- | --- | --- | --- |
| 18 | 9 Female/<br>9 Male | 11 Canada<br>4 Europe<br>1 Africa | 1 | Range: 1.5- 42<br>Mean: 17.5<br>Median: 17 | 38-78 | Rural only (n=7; 39%)<br>Urban only (n=5; 33%)<br>Both (n=6; 28%) | 9:<br>-long-term care<br>-military base<br>-hospitalist<br>-urgent care |
| Patient and Caregivers: | Gender | Age Range: | Ethnicity |  | Focus Group Length (minutes) | Disease represented |  |
| 8 bereaved caregivers<br>1 patient | 7 Females/<br>1 Male<br>1 Female | 40's-80's | 6 Caucasian<br>2 South East Asian<br>1 Caucasian |  | 90, 105, 118 | -lung cancer<br>-chronic lung disease<br>-colon cancer<br>-coronary artery disease<br>-blood cancer<br>-melanoma<br>-neurological cancer |  |
| Palliative Home Care Team Members: | Gender | Credential | Number per credential | Average years of clinical practice (years) | Focus Group Length |  |  |
| 26 | 21 females<br>5 females | RN | 17 | 17.5 (range 2-43) | 75 min<br>90 min |  |  |
|  |  | LPN | 2 | 11.5 (3, 20) |  |  |  |
|  |  | RRT | 2 | 11 (6, 20) |  |  |  |
|  |  | PT | 1 | 28 |  |  |  |
|  |  | SW | 3 | 21 |  |  |  |
|  |  | MD | 1 | 21 |  |  |  |

Distinct focus groups were used to discuss the perspectives and experiences of the patients, bereaved informal caregivers and PHC teams. Basic demographics such as age range, credentials, years of clinical practice, and medical condition represented were collected for these two groups (Table 1). A semi-structured focus group guide incorporating AI principles asked participants for their reactions and thoughts on the “Possibility Statements”[21] (Table 2) that were generated after analysis of the FP interviews in the first phase of the project (See Data Analysis for more details). Each of these specific groups had more than one focus group scheduled to accommodate the availability of consenting participants and were conducted by the principal author.

**Table 2:** Possibility Statements from Family Physician Appreciative Inquiry Interviews.

| **Possibility Statement** |
| --- |
| **1) Everyone (all clinicians, healthcare professionals, patients, caregivers, family) will value the longitudinal family physician-patient relationships with the patient as the focus.** |
| **2) All colleagues will consider the family physician as part of the <u>patient's</u> team.** |
| **3) There will be flexibility and nimbleness in system to respond to individual patient/family needs.** |
| **4) Each family physician will have a high-functioning team (within Family Medicine) to support the patient.** |
| **5) There will be enhanced multi-directional communication between *all clinicians* and care providers, leveraging technology as appropriate. The onus of the communication between clinicians must *not* fall on the patients and their caregivers.** |
| **6) Family physicians will champion the "Palliative Approach to Care" for their patients.** |
| **7) There will be ongoing, affordable and easily accessible educational opportunities for all healthcare providers who support patients with palliative care needs.** |
| **8) Remuneration for family physicians will be improved to better support the needed time (including travel time) and flexibility required to support patients in the community with palliative care needs ie: home visits, phone calls, team conferences virtually.** |

All interviews and focus groups were audio-recorded and transcribed into smart verbatim by an approved and confidential transcription service. Transcripts were checked for accuracy. Field notes capturing key observations from the interviews and focus groups were used to inform future interviews and focus groups as part of an iterative process.

## DATA ANALYSIS

### Family Physician Interviews

Qualitative analysis of the transcripts was conducted through line-by-line analysis for themes as they emerged by all three authors and a research assistant (SD). All three authors are experienced clinicians in palliative care as well as researchers, while the lead author also practices comprehensive family medicine. The research assistant did not have any clinical background. These varied perspectives facilitated grounding of the analysis in the real-life clinical context. The emerging themes were then used to develop a framework to which further analyses were compared to ensure that theme saturation was achieved through subsequent interviews [22] in an iterative manner. The research team members independently reviewed, analyzed the transcripts and met to discuss overall impressions regarding thematic content and to identify areas for further exploration in future stages of the data collection. Consensus was reached on the identified primary themes for the FP perspectives. These themes were then used to generate “Possibility Statements”[15, 21] according to AI methodology (Table 2). “Possibility Statements,” also known as “Provocative Statements,” [21] describe the ideal future destination in affirmative language and present tense without stipulating how to achieve this future destination.

### Focus Groups with Palliative Home Care Team Members, and Patients/Caregivers

The transcripts from the different focus groups were analyzed qualitatively using line-by-line analysis for themes as they emerged inductively by the principal author and a research assistant (CP). The focus group transcripts were also analyzed deductively [23] against the “Possibility Statements” that were derived from the family physicians to test the future destination from other perspectives.

### Development of Conceptual Framework to Describe the “Destiny” State

Consensus was reached by the 3 authors, through discussion of the themes as part of the iterative analysis, allowing for a deeper understanding of the different themes that had emerged. An audit trail kept throughout the project allowed for constant comparison of the data and memorandums were used to delve into the interpretation of the data, and of emerging relationships between themes and concepts. As these memos were sorted and grouped through constant comparison, relationships emerged that created categories that led to the development of the conceptual framework [24] for achieving the “Destiny” state.

### Rigour of Study Methods [25]

In keeping with accepted criteria for rigour in qualitative research (credibility, dependability, confirmability and transferability), our research team detailed the approach to analysis clearly and ensured we had appropriate expertise within our research team. The semi-structured interview guides were pilot-tested with similar test subjects who did not participate in the actual study. Purposeful sampling achieved an excellent variety of perspectives to ensure a well-rounded exploration in the interviews and focus groups.

Analysis was performed initially by the principal investigator (AT) and a research assistant (SD). Inter-coder reliability was ensured by AT and SD meeting at regular interviews and individually coding transcripts. The other two authors were also involved in individually coding the transcripts and regular meetings with the principal author. These iterative processes ensured coding accuracy, deepened the emerging understanding of the analysis and achieved consensus through discussions of differing views. Saturation was surpassed for all three groups that participated in interviews and focus groups. Memos and an audit trail were kept throughout the study from inception through to analysis to developmental of the conceptual framework. The analysis of the three groups of participants was completed separately and then triangulated with each other through constant comparison to create the thematic framework that laid the foundation for the conceptual framework.

The conceptual framework was triangulated with family physicians, caregivers, palliative care providers and other stakeholders who were not participants in the study to ensure that it resonated with their perspectives as an additional check for validity.

## RESULTS

### Demographics

The demographics collected for the three groups of participants for this study are detailed in Table 1 below.

12 of the family physicians self-identified as actively providing palliative care as part of their practice while 6 physicians did not include palliative care as a substantial portion of their practice. These 6 physicians did agree philosophically that primary palliative care was within the scope of a family physician to some extent but lacked practical experience in implementing this as part of their practice.

### Possibility Statements through Appreciative Inquiry Family Physician Interviews

The Appreciative Inquiry approach to the family physician interviews and analysis uncovered eight key areas. Focusing on these areas will better support them to improve CPPC for their patients. These eight themes created the Possibility Statements that described the family physician participants’ collective vision for a future “Destiny” state as part of the Appreciative Inquiry principles.

*“I think the holistic approach of family medicine, [is] of treating the patient, treating the family, treating the disease symptoms, and also …the whole being of the patient as well. So, the palliative process as well. And so they [family physicians] should, I think, have an important role in that*.*”*

### Results from the Focus Groups with Patients and Bereaved Caregivers and Palliative Home Care Team

The Appreciative Inquiry approach was used in the focus groups with both the Patients and Bereaved Caregivers and the Palliative Home Care Teams. The themes derived from the focus groups were triangulated with each other and are organized below in this table. Exemplar quotes to support these themes are found in the expanded table in Appendix 2.

### Conceptual Framework

Based on the themes described (Table 3) in an iterative manner [24], above, a conceptual framework emerged that brought all different perspectives together to describe how to achieve this “Destiny” state. This conceptual framework is shown in a visual schematic below with the symbols in the legend and describes how clinicians, the health system and society can more optimally help improve the patient’s and family’s journey when a person has palliative care needs. The following will describe the details that the visual schematic represents.

**Table 3:** **Themes from Focus Groups held with Family Physicians, Patients and Caregivers, and Palliative Home Care Teams**

| CATEGORY | Family Physicians (FP) Themes | Patient & Caregivers Themes | Palliative Home Care (PHC) Themes |
| --- | --- | --- | --- |
| 1) PATIENT'S RELATIONSHIP WITH FP NEEDS TO BE FOSTERED & VALUED | -Value of FP-patient relationships for the patient<br>-Colleagues to consider FPs part of the <u>patient's</u> team | -Encourage ongoing relationship with FP throughout illness so FPs have been in the loop<br>-FPs can help patients/caregivers navigate system & illness course | -Encourage ongoing relationship with FP throughout illness so FPs have been in the loop |
| 2) COMMUNICATION | -Enhanced Communication between care providers<br>-Telecommunication & person-to-person modes needed | -All of patient's team members communicate with *each other*, not through patient | -First point of contact b/w Home Care & FPs is crucial<br>-Improved, ongoing 2-way communication with FP using telecommunications more |
| 3) TEAM TO HELP SUPPORT PROVIDERS → IMPROVED SUPPORT FOR PATIENT/FAMILY | -Team-Based Care within FM team to support patient<br>-Team-Based Care with Palliative Home Care to support patient | -Psychosocial support is necessary for caregivers (beyond death) plus patients | -Palliative Home Care Manager Role: to advocate for & support patients/families |
|  |  |  | -Palliative Home Care Manager Role: can be eyes and ears for FP |
| 4)<br>UNDERSTANDING<br>PALLIATIVE<br>APPROACH TO<br>CARE | -FPs to champion Palliative Approach to Care | -Early discussion of what Palliative Care really is so can accept help & understand this in making treatment decisions (ACP and Goals of Care) | -Health care system & public need to better understand Palliative Care |
| 5) HEALTHCARE<br>SYSTEM NEEDS | -Need flexibility and nimbleness in system<br>-Remuneration for FPs needs to improve (travel, telecommunication visits) | -System should support patients in the community, *not rely on informal caregivers*<br>-More resources for different levels of home care needed throughout trajectory<br>-FPs need to be able to have longer appts<br>-Use telecommunication technology to be able to be in easier contact with FPs for questions, not necessarily home visits. | -Better transitions and handovers between care sectors<br>-More resources for different levels of home care needed throughout trajectory |
| 6) EDUCATION | -Ongoing educational opportunities for providers | -Public education about what palliative care is | -Ongoing educational opportunities for providers |

### Overall Conceptual Framework Description

The conceptual framework (Figure 1) places the patient with their family at the centre, surrounded by caregivers supporting the patient in the community. Alongside the patient and caregivers is the family physician within the Patient Medical Home as the longitudinal support that is the foundation of the Canadian healthcare system. The patient and caregivers’ journey is directly influenced by two inter-related relationship triads. We use the concept of the “**Loran”**[26], short for **LO**ng-**R**ange **A**id to **N**avigation, a system of navigation that uses signals between three points to determine one’s course for a journey, in this case the journey of the patient, family & caregivers. These relationship triads exist and interact with each other within the healthcare system and the larger society in which we all live. The key parts of the Loran conceptual framework are: 1) Two inter-relationship triads, 2) Triads exist within health system, and 3) Triads and health system exist within a larger society.

**Figure 1:**
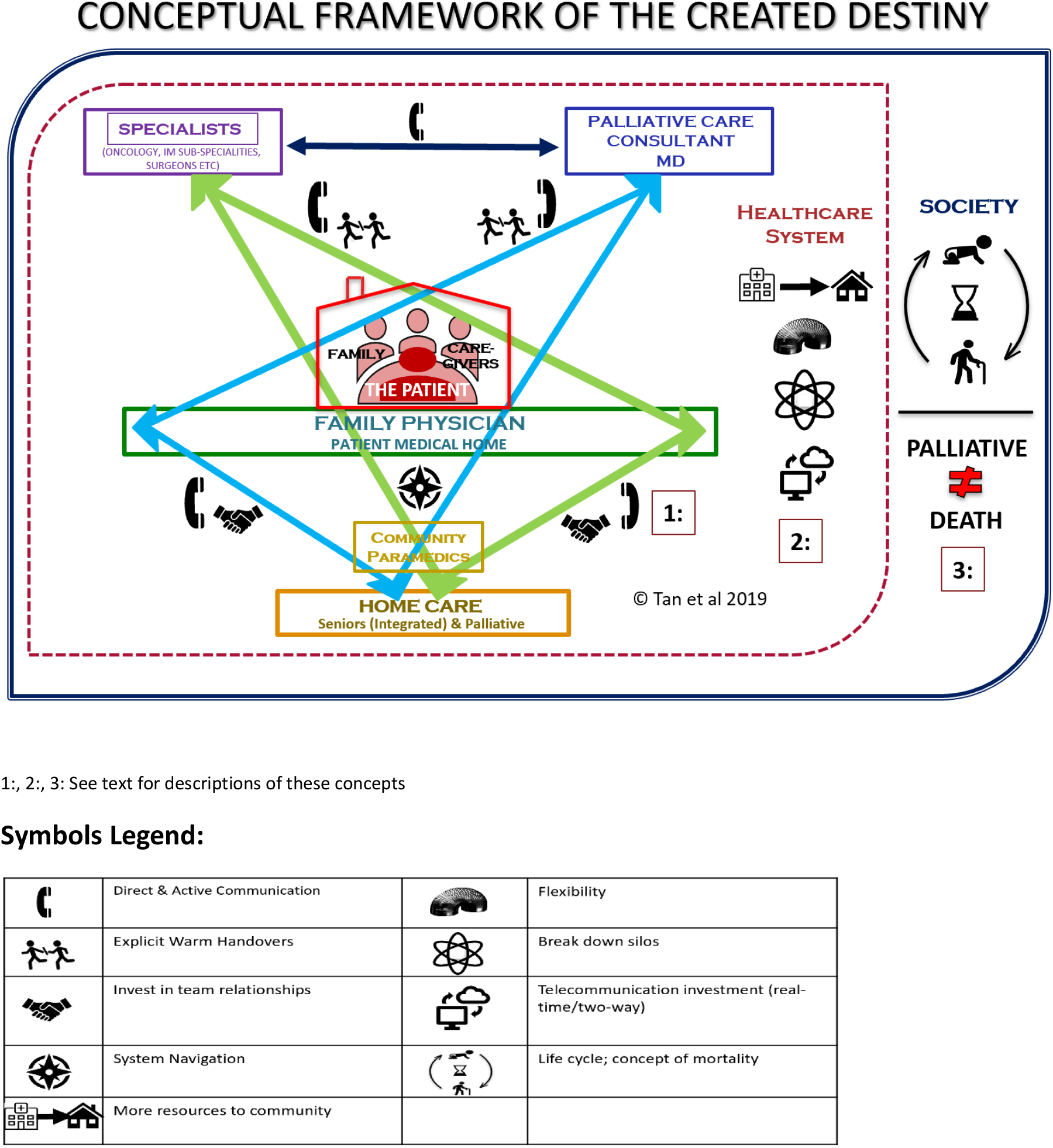
Conceptual Framework of the Panoramic View on how to Achieve the “Destiny” State.

### The Two Inter-Relationship Triads based on the “Loran” Concept [1]

The **first** relationship triad is between the patient’s family physician, home care providers and specialist consultants (e.g. oncologists, surgeons). The **second** relationship triad is between the patient’s family physician, home care providers and palliative care consultants, including advanced practice nurses, nurse practitioners and consultant physicians.

Our analysis found that these relationship triads are positively impacted by the *upstream enablers* that each clinician would have enacted prior to working within the triad. These upstream enablers optimized the possibility of an effective, collaborative working relationship between these clinicians to improve the patient’s journey together. Once working together, the framework also illustrates the ongoing *facilitators* that emerged as essential to enhance the collaboration amongst the clinicians working with the patient and caregivers.

**Figure 2:**
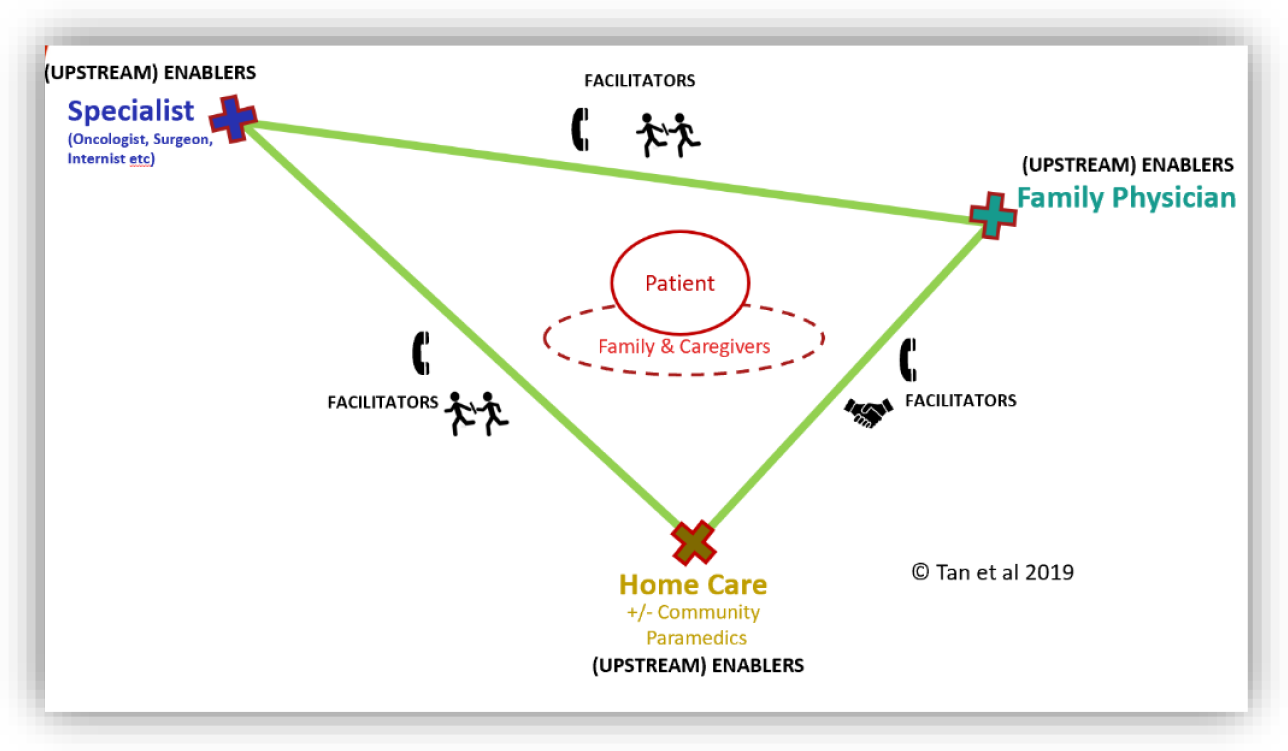
First Relationship “Loran” Triad: Family Physician – Home Care – Specialist Consultant(s)

**Figure 3:**
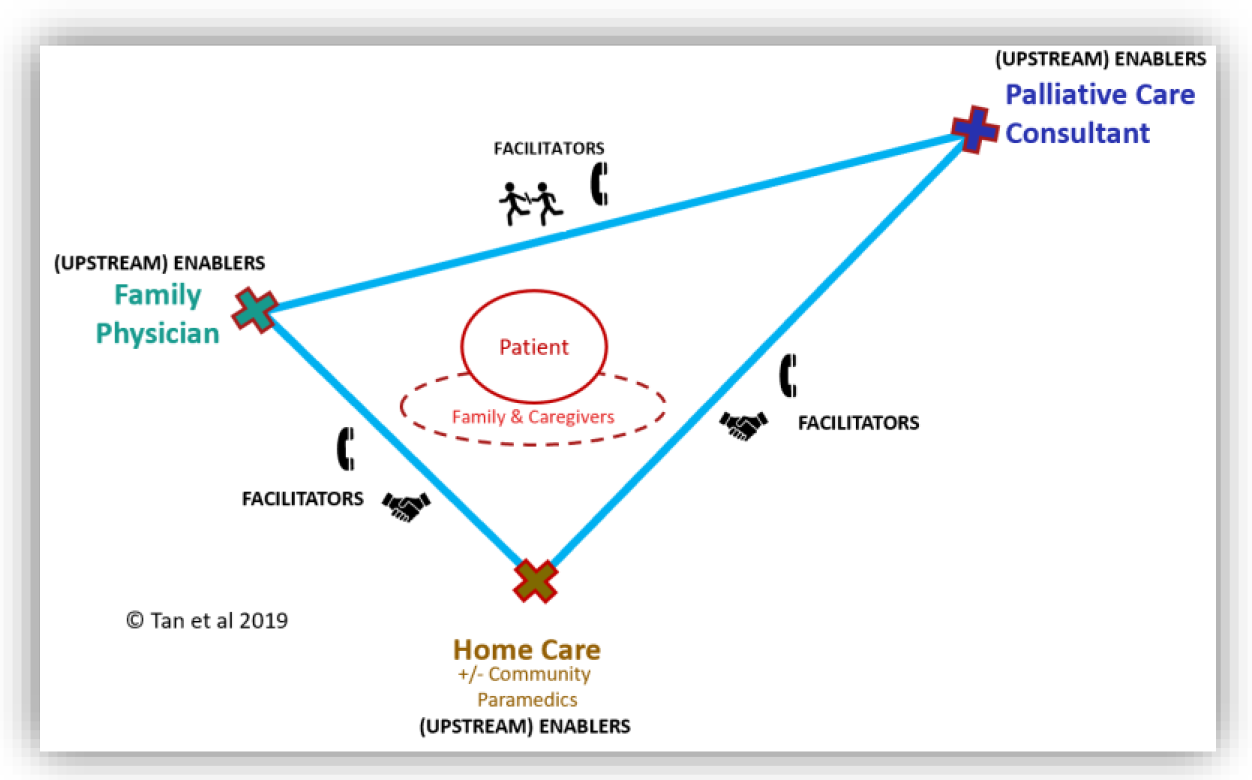
Second Relationship “Loran” Triad: Family Physician – Home Care – Palliative Care Consultant(s):

**Figure 4:**
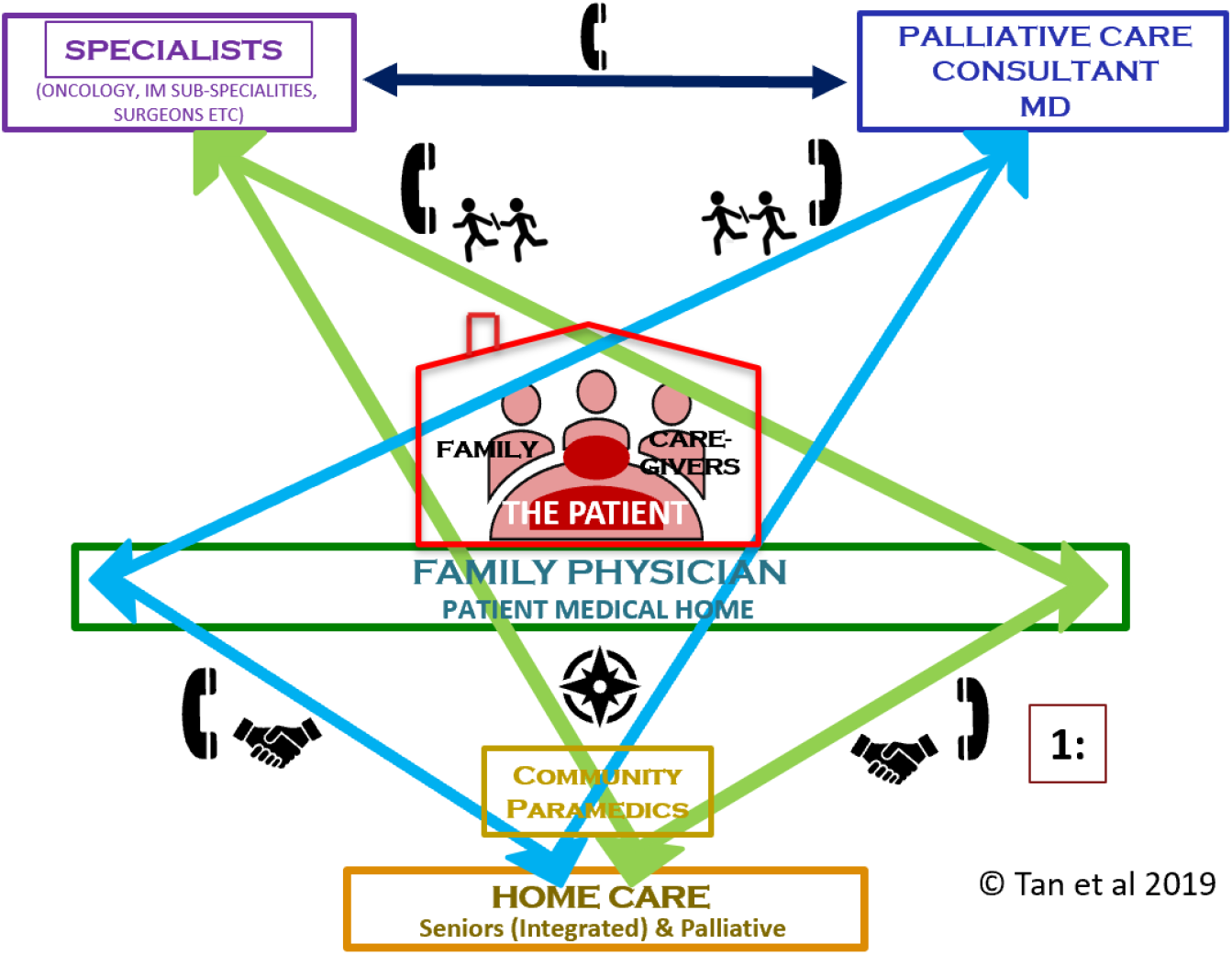
Both “Loran” Triads Overlapping with Upstream Enablers and Facilitators Incorporated.

Table 4 details the upstream enablers and the facilitators that would enhance collaboration amongst the clinicians, for the benefit of the patient and caregivers.

**Table 4:**
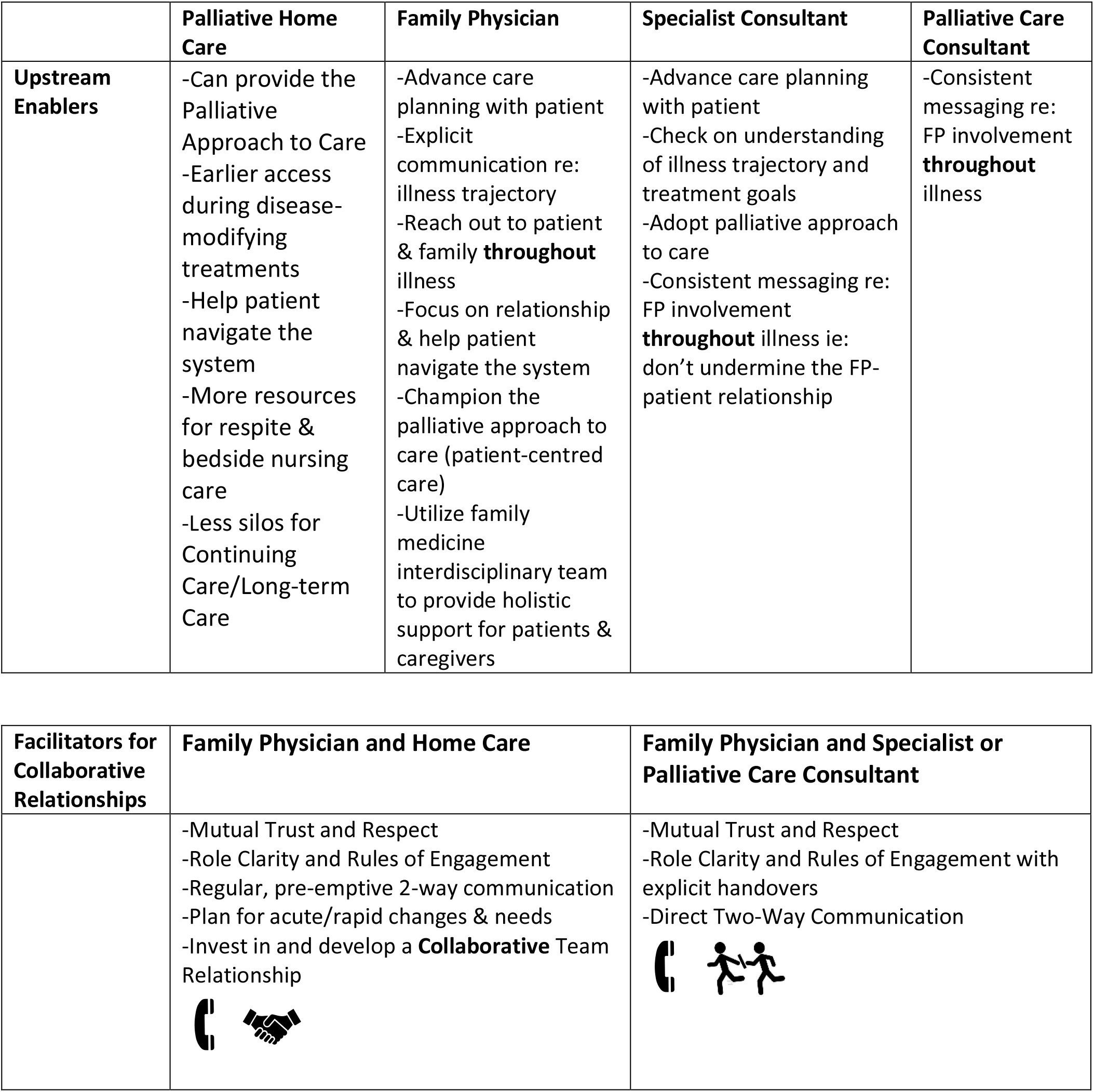
**The Upstream Enablers Enhancing Collaboration among Clinicians for care of Patients and Caregivers**

|  | Palliative Home Care | Family Physician | Specialist Consultant | Palliative Care Consultant |
| --- | --- | --- | --- | --- |
| **Upstream Enablers** | -Can provide the Palliative Approach to Care<br>-Earlier access during disease-modifying treatments<br>-Help patient navigate the system<br>-More resources for respite & bedside nursing care<br>-Less silos for Continuing Care/Long-term Care | -Advance care planning with patient<br>-Explicit communication re: illness trajectory<br>-Reach out to patient & family **throughout** illness<br>-Focus on relationship & help patient navigate the system<br>-Champion the palliative approach to care (patient-centred care)<br>-Utilize family medicine interdisciplinary team to provide holistic support for patients & caregivers | -Advance care planning with patient<br>-Check on understanding of illness trajectory and treatment goals<br>-Adopt palliative approach to care<br>-Consistent messaging re: FP involvement **throughout** illness ie: don't undermine the FP-patient relationship | -Consistent messaging re: FP involvement **throughout** illness |

| Facilitators for Collaborative Relationships | Family Physician and Home Care | Family Physician and Specialist or Palliative Care Consultant |
| --- | --- | --- |
|  | -Mutual Trust and Respect<br>-Role Clarity and Rules of Engagement<br>-Regular, pre-emptive 2-way communication<br>-Plan for acute/rapid changes & needs<br>-Invest in and develop a **Collaborative** Team Relationship | -Mutual Trust and Respect<br>-Role Clarity and Rules of Engagement with explicit handovers<br>-Direct Two-Way Communication |

### The Loran Triads exist within a Healthcare System <u>(dotted red line</u>) [2]

The conceptual framework is grounded by these two Loran triads that influence the patient and caregiver illness journey. However, even if these Loran triads are well established with all clinicians having engaged in the upstream enablers listed in Table 4, and working collaboratively together, it must be acknowledged that these cannot exist in isolation. These relationships must exist in the healthcare system, which also requires improvements to better support patients and caregivers in their community-based palliative care journey.

**Figure 5:**
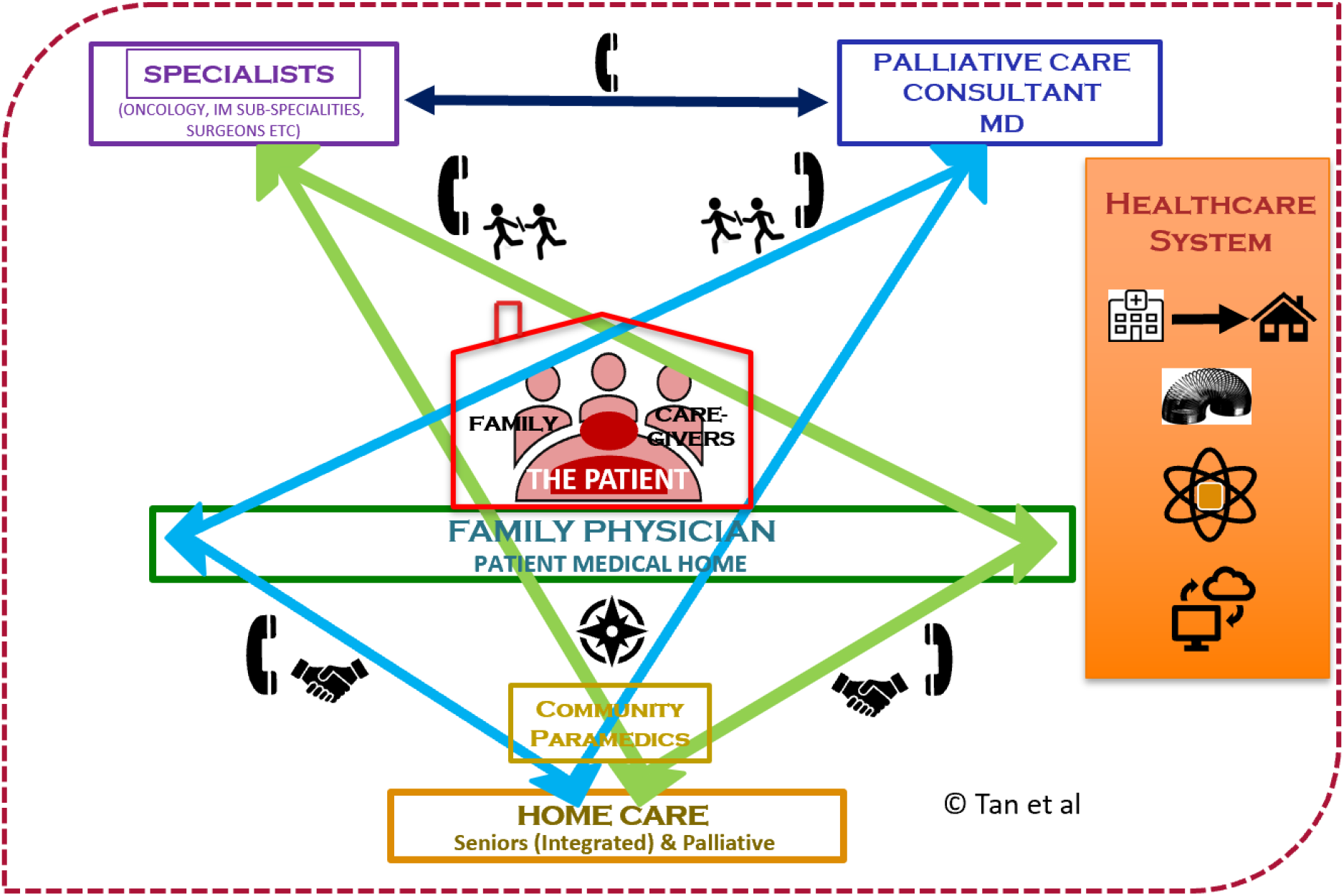
The Loran Triads exist within a Healthcare System <u>(dotted red line</u>)

The key components that were identified as required within the healthcare system were: 1) resources focused more on community than acute care facilities, 2) system flexibility to respond to individual patient needs, 3) elimination of silos between clinicians, disciplines and organizations to improve collaboration and efficiency and decrease redundancies and confusion, 4) use of secure teleconferencing modalities to improve access and support for patients regardless of location, and 5) a universal electronic record system to improve communication amongst all clinicians, care providers and patients.

In turn, the healthcare system exists within the larger society that it serves.

**Fig 6:**
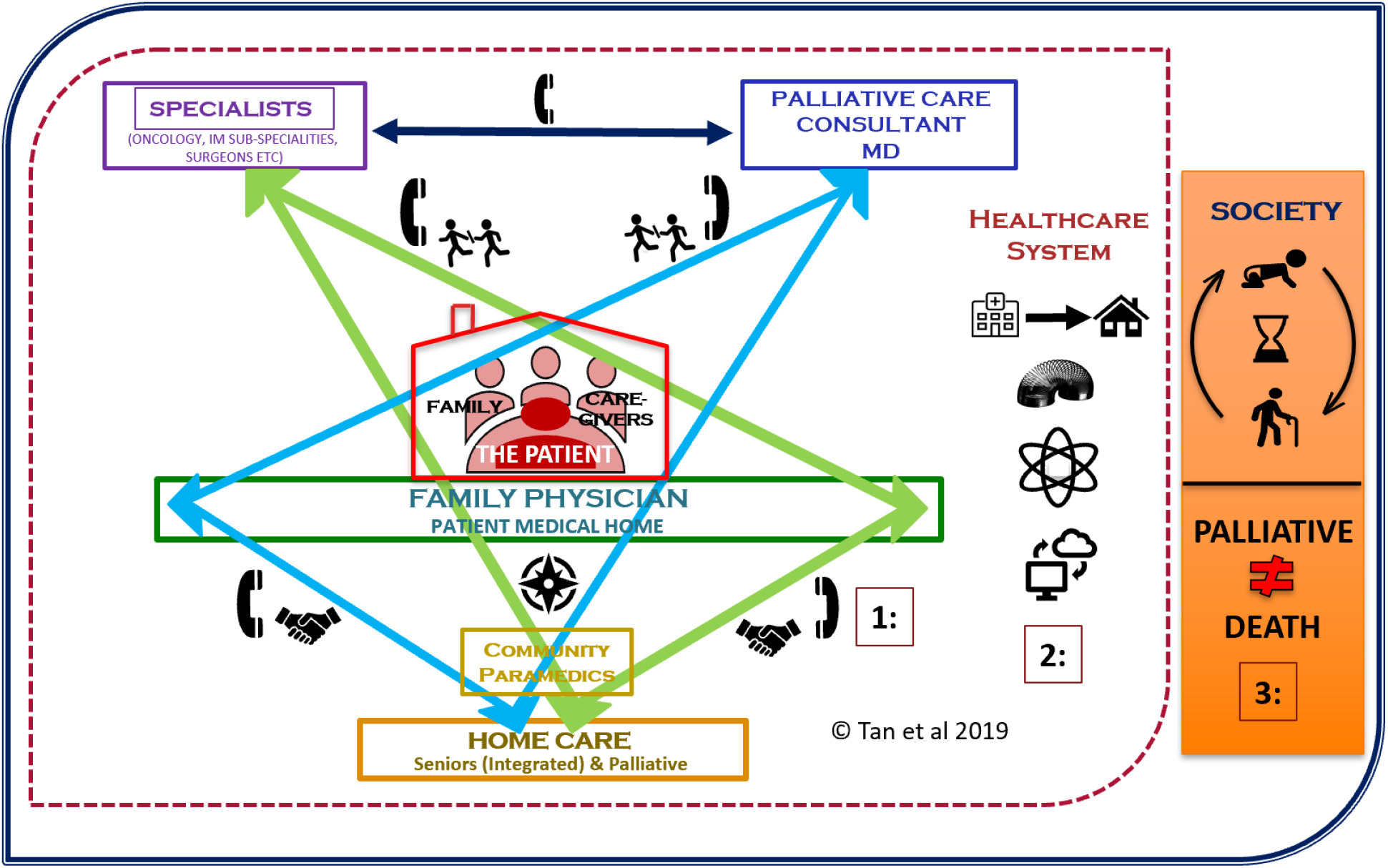
The Healthcare System & the Loran Triads exist within the Larger Society (outer blue line) [3].

Analysis identified that patients, caregivers, and clinicians agreed that more public education is required to help everyone inside and outside the healthcare system understand that Palliative Care does ***not*** mean end of life or imminent death. This would allow involvement of palliative care supports earlier in the disease trajectory. Additionally, public discourse will normalize conversations about mortality and decrease stigma and fears about death, dying and appropriate treatment decisions in accordance with patients’ goals.

## DISCUSSION

This study explores and describes behaviours and health system components that are required to improve the coordinated and collaborative care for community-based patients with palliative care needs and their family and unpaid caregivers. We interviewed FPs who practice in a large Canadian city and surrounding rural area. Using AI methodology, we identified “Possibility Statements” that described a future “Destiny” state. These “Possibility Statements” were presented to bereaved caregivers and patient focus groups, as well as two PHC teams, to determine their reactions and perspectives to then create a panoramic view of the “Destiny” state. Based on our analysis, we developed a conceptual framework that describes how to achieve this “Destiny” state with the patient and family journey as the central focus. It is anchored by two “Loran” triads that describe the effective collaboration required between the four different groups of clinicians (FPs, specialists, palliative care consultants, home care), who support the patient and family.

This conceptual framework is a significant development in the primary palliative care literature as its key contribution identifies the explicit ways that the healthcare team, healthcare system and societal attitudes can be optimized to improve the care of patients with palliative care needs. While the roles and responsibilities of different healthcare providers have been explored in the provision of palliative care [5, 9], this is the first comprehensive framework that pulls all of these perspectives and roles together.

Many identified themes transcended all three interviewed groups. These themes created the foundation for the conceptual framework. Enhanced communication and the fostering of relationships among all care providers, with the focus on the patient and caregivers, is the cornerstone concept.

Encouraging strong longitudinal FP/patient relationships is essential. Increased system flexibility and improved home care resources are constant themes. Furthermore, patients and caregivers need more education regarding the benefits of palliative care, with increased public discourse about mortality.

Our study identified several other key findings that other studies have also reported. Effective teamwork amongst clinicians and care providers of all disciplines was universally recognized as the most important component [27-29]. The different perspectives elicited multiple ways to improve and optimize the patient and caregiver journey through the palliative care illness trajectory. With the patient as the central focus of the team [27-29], all care is guided by the patient’s preferences, values and needs [30].

Another key enabler identified was strong, continuous relationships between FPs and patients, focussing on the “whole person” [31] using the PMH [30] model throughout their illness, enhances care and avoids gaps in treatment. Continuous FP and PMH involvement can also help family members and loved ones access bereavement and grief support, reducing the existing unmet need where care ends once a patient is deceased [32]. PMHs have demonstrated success in improving access to the health system, enhancing quality of care, improving coordination of care, reducing reliance on acute care facilities and encouraging a team-based care model [33]. Family physicians within these PMHs can utilize the multidisciplinary team-based care model, reducing reliance on acute care facilities by patients [33-37] and encouraging a team-based care model.

Increased investment in community-based home care teams that are skilled in both the palliative approach to care and primary palliative care skills is essential to support patients in the community [38]. More home care resources for patients with palliative care needs and improved collaboration with family physicians lessens the burden for families. Studies have found that patients with palliative care needs desired regular home visits by their FP [39], yet our study found that patients and caregivers want ongoing availability of their FP on an *as-needed basis by phone* rather than regular or frequent home visits. A recent study determined that patients in rural communities found that the use of web-based teleconferencing with a physician consultant was a convenient and acceptable way to address their concerns and the physicians noted improved patient access in a time-efficient manner [40]. The home care team member played an essential bridge that supplied the teleconference equipment to the patient’s home, and also acted as the eyes and ears for the physician. The use of technology shows promise in encouraging more FPs to maintain regular contact with patients with palliative care needs in the community. While this study was conducted prior to the SARS-COV-2 pandemic, the sudden necessity to employ virtual care at the height of the Canadian public health lockdowns in March 2020 demonstrated the importance of the use of technology to support patient care [41, 42]. The lessons learned during the pandemic will expedite the sustained healthcare system changes needed to better support house-bound patients with palliative care needs beyond the pandemic [43]. Care must be taken to ensure virtual care *improves* continuity between care providers and the patient with palliative care needs, rather than allowing for convenience with episodic care to dominate over longitudinal relationships with a family physician.

The longitudinal relationship between a patient and trusted family physician must be encouraged, valued and fostered by everyone in in the care team. Strong collaboration builds respect, trust and competence among family physicians, the home care team and specialists [44]. Our study found similar enablers to improving team function for the patient’s benefit. Safe and open communication among team members in a supportive team environment, a clear understanding of team member roles, responsibilities and processes and providing space for flexibility resonated with all stakeholders who participated in our study. Nancarrow et al.[28] identified these features as the key principles required for good interdisciplinary team function. Unfortunately, the bereaved caregivers and patients in our study commented that they often received contradictory messages from specialists and palliative care consultants that undermined their relationship with and trust in their FP. An Ontario study found that patients with palliative care needs who had FP visits less than 6 months from death were facilitated by their satisfaction in their FP’s care, which included the desire for psychosocial support by their FP and caregiver support [45]. Affirmation of the role of the FP by all care providers and improved role clarity will improve trust and strengthen relationships between clinicians and care providers [45].

Family physicians pointed out that they desire to provide palliative care to their patients, but because these patients make up a small subset of their overall generalist practice, they need the support of the team. This includes easier access to their specialist colleagues for help or advice for their patients. Anvik et al. [31] also found that patient care improves when generalists and specialists reinforce each other. When specialists and family physicians can actively work together and share the care of their mutual patient, there is less risk of family physicians being left to “hold the bag” [27], and a collaborative “sharing of the load” [27] occurs that benefits the patient. Coaching family physicians to manage more complex palliative care situations will help to meet patients’ needs.

This study reiterates the concern that the medical community and society at large do not have a good understanding of what palliative care can offer patients and their family members. Palliative care *does not* mean that the patient is imminently dying. As clinicians improve their skills and embrace the palliative approach [46-48] to care, patients and families will be better supported in their serious illness journey and will be less threatened by the palliative approach to care. Open conversations about a patient’s illness trajectory and prognosis will lead to earlier and more effective advance care planning for patients [49]. Collaboration and improved communication between specialists and family physicians will result in less conflict in messaging and will help to ensure that caregivers are better [50] prepared for substitute decision-making. Increasing public discourse [51] about death and dying may empower clinicians to have important conversations with patients and might limit the use of ineffective or unwanted medical treatments. Compassionate and coordinated care in the PMH will reduce fear and improve conversations and decision-making in the midst of life-limiting illness.

### Strengths and Limitations of Study

This study’s strength lies in the innovative solutions-based Appreciative Inquiry approach to determine how best to support patients and their families in the community with palliative care needs.

Additionally, exploring the perspectives of family physicians, palliative home care providers and their patients and unpaid caregivers and triangulating their insights allowed for the development of a conceptual framework from important yet varied stakeholder perspectives.

As with any qualitative study, the results of this study may not be generalizable to other regions. The recruitment of patients and caregivers was limited in number and in diversity of demographics as we were dependent on volunteers through an arms-length organization for ethical reasons. However, the content of the focus groups involving patients and caregivers did surpass saturation in analysis and we were satisfied with the richness of the data collected for this perspective.

## Future Work

Future work would include implementation of the practical aspects of the conceptual framework that this study created that integrates with emerging best practices for improving purposeful team collaboration [5]. Studying the work of an initial small group of early adopters who learn effective team strategies, strengthen the homecare system and promote societal changes required to support patients with palliative care needs could inform others in implementation. More work is required to explore family physicians’ definitions, attitudes and comfort with integrating different components of palliative care. Identifying family physicians who are “palliphilic” (versus “palliphobic” [52]) could allow for these early adopters to implement the practical aspects in our conceptual framework toto promote sustainable system change.

## CONCLUSION

A strong, effective collaborative relationship between the family physician with all other care providers is critical to support patients with palliative care needs and their caregivers in the community, as is building on the trusting FP-patient longitudinal relationship. Key areas were identified for how all members of the patient’s team can work together effectively to improve the patient and caregiver palliative care journey.

## Data Availability

Data is stored locally on a secure harddrive and can be provided at request.

## Acknowledgements

The authors would like to thank Dr. Jessica Simon and Dr. Patricia Biondo for their help in the background work for this project. We would like to Sharlette Dunn and Nicole Frenette for this help in the data collection and analysis and formatting of the manuscript.

## Contributors

AT, RS and AS conceptualized the study. AT designed the study with feedback from RS and AS. AT led the data collection with the research assistant. All authors reviewed the transcripts and contributed to the analysis. AT led the manuscript writing with contributions from RS and AS. All authors reviewed the transcript.

## Funding

This work was supported by a Janus Research Grant from the Foundation for Advancing Family Medicine of the College of Family Physicians of Canada and a Grant from Alberta Health.

## Competing Interests

None declared.

## Patient consent for publication

Not required.

---

**Appendix 1:** **Detailed Description of Possibility Statements derived from Family Physician Appreciative Inquiry Interviews**

| Possibility Statement | Description |
| --- | --- |
| 1) Everyone (All clinicians, healthcare professionals, patients, caregivers, family) will value the longitudinal family physician-patient relationships with the patient as the focus. | -more education and understanding about the family physician's central role in a patient's care as a care coordinator of any/all health concerns while providing continuity of care throughout a longitudinal relationship.<br>-acknowledging and understanding the broad scope of a family physician's practice including supporting patient's family/unpaid caregivers throughout the illness and into bereavement, if necessary. |
| 2) All colleagues will consider the family physician as part of the <u>patient's</u> team. | - regardless of where a patient is receiving care in the healthcare system, a patient never stops being their family physician's patient.<br>- FP's are consistently part of the patient's care team; when medical colleagues include them in care planning, discharge planning and ongoing care of their mutual patients, the patient and their family benefit from coordinated and collaborative care.<br>- investing in relationships with these patient team members will improve care for patients. |
| 3) There will be flexibility and nimbleness in system to respond to individual patient/family needs. | -healthcare system needs to allow for more flexibility to more nimbly respond to the unique needs and challenges that individual patients and families face. |
| 4) Each family physician will have a high-functioning team (within Family Medicine) to support the patient along with the family physician. | - having multidisciplinary team support within their individual family medicine practice is an essential and valuable support in meeting the needs of patients with palliative care in their practices.<br>- access to team members as nursing staff, clinical pharmacists, social workers helps FPs to more effectively support patients and families and better meet their needs. |
| 5) **There will be enhanced multi-directional communication between *all clinicians and* care providers, leveraging technology as appropriate. The onus of the communication between clinicians must *not* fall on the patients and their caregivers.** | - includes the use of technology such as secured messaging or interactive electronic medical records to facilitate two-way communication.<br>- be purposeful to carve out time through booked teleconference calls between specialists and family physicians, or home care providers and family physicians to discuss shared care of patients.<br>-collaboratively manage concerns and/or pre-emptively managing inevitable changes. |
| 6) **Family physicians will champion the “Palliative Approach to Care” for their patients.** | -leverage FPs to champion the Palliative Approach to Care at the appropriate times in the patient’s illness trajectory.<br>-FP can use the rapport built from the ongoing relationship with the patient, and the medical knowledge of the patient’s context to provide support and education about illness trajectories, treatment options and overall goals of care that could guide decision-making with the patient and family. |
| 7) **There will be ongoing, affordable and easily accessible educational opportunities for all healthcare providers who support patients with palliative care needs.** | -ongoing support and educational opportunities are needed to support FPs who have different comfort and experience levels with providing support to patients with palliative care needs within their scope as a family physician. |
| 8) **Remuneration for family physicians will be improved to better support the needed time (including travel time) and flexibility required to support patients in the community with palliative care needs ie: home visits, phone calls, team conferences virtually.** | - FPs must be compensated for travel time and taking time away from their clinics to maximally supporting patients in the community with palliative care needs, especially if their clinical condition were to deteriorate so that they are more house-bound. |

**Appendix 2:** **Themes with Exemplar Quotes from Focus Groups held with Family Physicians, Patients and Caregivers, and Palliative Home Care Teams (Expansion of Table 3)**

| CATEGORY | Family Physicians (FP) Themes | Patient & Caregivers Themes | Palliative Home Care (PHC) Themes |
| --- | --- | --- | --- |
| **1) PATIENT'S RELATIONSHIP WITH FP NEEDS TO BE FOSTERED & VALUED** | -Value of FP-patient relationships for the patient<br>-Colleagues to consider FPs part of the <u>patient's</u> team | -Encourage ongoing relationship with FP throughout illness so FPs have been in the loop<br>-FPs can help patients/caregivers navigate system & illness course | -Encourage ongoing relationship with FP throughout illness so FPs have been in the loop |
| *Exemplar quotes:* | *"And again...the relationship they've developed with their family doctor. And so, just make it easier to talk about these then...maybe they're more comfortable with bringing up their concerns about dying, or concerns about pain and suffering at the end of life. And concerns about taking care of the loved one, any of those things that would be easier to talk to, about the doctor you've known for 20 years or something." FP 1, male* | *"the [family doctor said] I think the time has come.... And I think it sort of sent home to me that the role of the family physician is so important to work with the palliative care team and the patient."-Caregiver FG 1*<br>*"[F]P's should communicate – basically level set, "I know what you're going through. And by the way this is my role for patients going through this type of disease." Pt/Caregiver FG 3, male* | *"there's the ones where a family member goes into the [FP] to keep up the relationship, and then the [FP] will call us and say, "This is what they're asking for. Is it appropriate?" "it's just somebody cares, they actually care enough to engage. And then what I find is then the clients are very protective of that relationship, they are very adamant you do not cut my physician out" PHC, female* |
| **2) COMMUNICATION** | -Enhanced Communication between care providers<br>- Telecommunication & person-to-person modes needed | -All of patient's team members communicate with *each other*, not through patient | -First point of contact b/w Home Care & FPs is crucial<br>-Improved, ongoing 2-way communication with FP using telecommunications more |
| Exemplar quotes: | *"if the patients visited them, identified new issues, changed meds , you had changed strategy... I'd like to hear from- from them probably more proactively to feedback to me.... I probably looked after the patient for years before that, so I kind of like to be kept maybe more in the loop from the palliative care team if I was to say anything"*<br>FP 9, male | *"Here you had to broker these. It's all fragmented... And nobody's talking to each other, so you have to actually remove the conflicts. because even that we had to do the scheduling. The scheduling conflict was on me."* Caregiver, F | *"you know you go to any meeting and there's people calling in so I'm sure they're doing it at other health care points, so maybe they [the FP] could do that, come into our rounds, especially for the really complex patients."* PHC FG 2, female |
| **3) TEAM TO HELP SUPPORT PROVIDERS → IMPROVED SUPPORT FOR PATIENT/FAMILY** | **-Team-Based Care within FM team to support patient**<br>**-Team-Based Care with Palliative Home Care to support patient** | **-Psychosocial support is necessary for caregivers (beyond death) plus patients** | **-Palliative Home Care Manager Role: to advocate for & support patients/families**<br>**-Palliative Home Care Manager Role: can be eyes and ears for FP** |
| Exemplar quotes: | *"co-manage patients with somebody they know"* FP 3, male<br>*"teams work because they've worked together".* FP 5, female | *"I personally, at that time, actually needed a bit more support than I was getting. I tried to seek it out, and didn't find it."*<br>Pt/Caregiver FG 1, female<br>*"So a possible improvement is to consult with a caregiver and say, "How can we help you help your loved one?" So that would be a route that I didn't see taken"*<br>Caregiver FG 2, female | *"To be an advocate for the patient. And just to be the eyes and ears, right? For the patient. Yeah, and the family. And also to support the doctor if the doctor needs some support."*<br>PHC FG 1, female 1 |
| **4) UNDERSTANDING PALLIATIVE APPROACH TO CARE** | **-FPs to champion Palliative Approach to Care** | **-Early discussion of what Palliative Care really is so can accept help & understand this in making treatment decisions (ACP and Goals of Care)** | **-Health care system & public need to better understand Palliative Care** |
| Exemplar quotes: | *"My job is to try to help them choose the best possible treatment for them"* | *"It's terminal, so you're going to go in palliative care and that's where the death, death, death, death, I* | *"That can be hit and miss depending on the physician's perception of palliative care, where they* |
|  | *with what we know. I'm a partner with the patient. And an advisor. I think what I can do is focus more on the individual. I can say let's talk about what might happen if this happened to you and your family; which path do you want to take. And it's rewarding, in the sense that I am able to provide counsel."* | *don't want that."* Pt/Caregiver FG 1, female<br>*"For them they're telling me it's palliative care. There's not a lot of options. And I didn't accept that."* Pt/Caregiver FG 3, male<br>*"He [My oncologist] said, and if your treatment limits your enjoyment of life what the heck is the whole point, eh? It's kind of along the lines we can do lots of things to you, but I want to do things for you."* Patient FG 1 | *just believe [unintelligible]. That can be tricky, as well, because I think we have conversations where you're like, "You studied medicine?" Like, it's weird. We're having this conversation that you don't – like, he literally did not want to talk about palliative care. This is a weird conversation."* |
| **5) HEALTHCARE SYSTEM NEEDS** | **-Need flexibility and nimbleness in system**<br>**-Remuneration for FPs needs to improve (travel, telecommunication visits)** | **-System should support patients in the community, *not rely on informal caregivers***<br>**-More resources for different levels of home care needed throughout trajectory**<br>**-FPs need to be able to have longer appts**<br>**-Use telecommunication technology to be able to be in easier contact with FPs for questions, not necessarily home visits.** | **-Better transitions and handovers between care sectors**<br>**-More resources for different levels of home care needed throughout trajectory** |
| *Exemplar quotes:* | *"And if somebody wants to be cared from home, and it would be beneficial to the patient to have their family doctor see them at home, it's not a great compensation for doing that. And especially in a city where you might have a patient that lives in the south end, or the north end, and it takes you an hour to drive there and then* | *"The expectation of the medical system...is high [on family/informal caregivers]. I have a medical background and found it hard. I don't know what people do if this is all new to them."* Caregiver FG 2, female<br>*"And it was all or nothing, and I think that would be probably my biggest problem with the service. Because I think at different stages of terminal cancer you need more services, and then you need less service."* Caregiver FG 1, female<br>*"So perhaps even in the billing,* | *"if we get the patients earlier in their trajectory we have more time to engage the [FP] as opposed to if we get them when they're already in crisis" PHC FG 2*<br>*"The average length of stay is one day till less than four months. And they're very sick and they're complicated. So if we're trying to find a new family doctor when we're looking at a one-month prognosis, it becomes very hard. We're* |
|  | *you don't get paid for your travel time. You don't get paid for this, and then it's the equivalent to getting paid for 15 minutes seeing a patient in the ...So if there's a way to compensate doctors for doing the home visits specifically for palliative care, taking into account travel time out in the field."*<br>FP 1, male | *you know, how they bill, when you've got a palliative care patient, perhaps they [the FP] can bill a little bit longer to give them a half an hour that they might needs."*<br>FG 2, caregiver female | *too late to the party already. We didn't start palliative care early enough. And so if we could shift everything earlier – that I think we can develop relationships."* |
| **6) EDUCATION** | **-Ongoing educational opportunities for providers** | **-Public education about what palliative care is** | **-Ongoing educational opportunities for providers** |
| *Exemplar quotes:* |  | *"when you were told that well by this rude homecare person that he was palliative since day one, would it make a difference if the word was different? Like if you were offered that support from a family doctor without using the palliative word, just because I know that palliative care can start from day one. But people think death as soon as they hear it."*<br>*"we thought palliative care was end-of-life care. And I think that's a common misconception."* |  |

## Notes

### Competing Interest Statement

The authors have declared no competing interest.

### Author Declarations

Health Research Ethics Board Approval: University of Calgary: REB17-1230

